# A Systematic Review evaluating the role of the 7 T MRI in Neurosurgery: Unveiling Trends in the Literature So Far after Clinical Approval in 2017

**DOI:** 10.1101/2023.11.12.23298426

**Authors:** Arosh S. Perera Molligoda Arachchige

## Abstract

**Aims:** (1) To assess the impact of 7-Tesla Magnetic Resonance Imaging (7T MRI) on neurosurgery, focusing on its applications in diagnosis, treatment planning, and post-operative assessment. (2) To systematically analyze and identify patterns and trends in the existing literature related to the utilization of 7T MRI in neurosurgical contexts.

**Methods:** Systematic search of PubMed for studies published between January 1, 2017, and December 31, 2021, using MeSH terms related to 7T MRI and neurosurgery. Inclusion criteria: Studies involving patients of all ages, meta-analyses, systematic reviews, and original research. Exclusion criteria: Pre-prints, studies with insufficient data eg: case reports, letters, etc., non-English publications, and studies involving animal subjects. Data synthesis involved standardized extraction forms, and a narrative synthesis was performed.

**Results:** Identified 83 records from PubMed, screened for inclusion criteria, resulting in 48 studies included in the systematic review. Most common neurosurgical procedures using 7T MRI: endoscopic neurosurgery, resective epilepsy surgery, and deep brain stimulation surgery. Commonly treated pathologies: cerebrovascular diseases, epilepsy, pituitary adenoma, and gliomas. USA, Netherlands, and Germany had the highest absolute number of publications, but the Netherlands showed the highest research productivity per 7T MRI available. A decline in research output in 2018 followed by an increase in subsequent years. Could not conduct a meta-analysis due to insufficient studies for each pathology.

**Conclusions:** 7T MRI holds great potential in improving the characterization and understanding of various neurological and psychiatric conditions. Superiority of 7T MRI over lower field strengths demonstrated in terms of image quality, lesion detection, and tissue characterization. Applications in epilepsy, pituitary adenoma, Parkinson’s disease, cerebrovascular diseases, trigeminal neuralgia, traumatic head injury, multiple sclerosis, glioma, and psychiatric disorders. Limitations include database selection, research productivity metrics, and study inclusion criteria. Findings suggest the need for accelerated global distribution of 7T MR systems and increased training for radiologists to ensure safe and effective integration into routine clinical practice.

## 1. Introduction

In the realm of modern medicine, the field of neurosurgery stands as a testament to the remarkable progress achieved through advancements in technology and imaging. The delicate and intricate nature of the human brain necessitates tools and techniques that allow for precise diagnosis, treatment planning, and surgical intervention. Among the many innovations that have revolutionized the field, one technology has emerged as a promising game-changer: the 7-Tesla Magnetic Resonance Imaging (7T MRI) scanner, which also received US FDA approval for clinical use in 2017 [1,2,3]. The introduction of 7T MRI represents a pivotal moment in the history of neurosurgery. This remarkable imaging technology harnesses the power of ultra-high magnetic fields to produce images with unprecedented detail and resolution, far surpassing the capabilities of conventional MRI scanners [4]. By capitalizing on the inherent magnetic properties of hydrogen nuclei within the human body, 7T MRI offers both neurosurgeons and neurologists an invaluable tool to investigate deeper into the intricate structures of the brain, enabling them to make more precise assessments of pathology and formulate optimized treatment strategies [5,6].

Traditionally, neurosurgery has relied upon lower field strength MRI machines, typically operating at 1.5 or 3 Tesla. While these systems have been instrumental in guiding surgical interventions and aiding in preoperative planning, they often fall short in providing the level of detail required for complex neurosurgical cases [2,3]. The limitations of lower field strength MRI, such as reduced spatial resolution and limited contrast, have posed challenges in accurately delineating critical structures, identifying subtle abnormalities, and characterizing lesions, ultimately affecting the quality and safety of neurosurgical procedures [5,6]. The emergence of 7T MRI, with its higher magnetic field strength, has raised the bar for neuroimaging capabilities. It promises to unveil new dimensions of information that were previously concealed, shedding light on intricate anatomical details and subtle pathologies that were once elusive [5,6]. This newfound precision has the potential to redefine the landscape of neurosurgery by offering enhanced preoperative assessment and surgical guidance, ultimately leading to improved patient outcomes and reduced surgical risks.

This systematic review aims to systematically analyze and identify patterns and trends within the existing literature on the utilization of 7T MRI in neurosurgical contexts, focusing on its applications in the diagnosis, treatment planning, and post-operative assessment of various neurosurgically treated pathologies.

## 2. Methods

A systematic search of the PubMed database was conducted to identify relevant studies published between January 1, 2017, and December 31, 2021, using the following MeSH search terms: ((7 Tesla MRI) AND (neurosurgery)). Two independent reviewers performed the initial search and screening, with inclusion criteria encompassing studies involving patients of all ages (both paediatric and adults) where 7-Tesla MRI imaging was performed and/or compared with conventional MRI imaging in the context of neurosurgery. Eligible study types included meta-analyses, systematic reviews, and original research. Exclusion criteria involved studies with insufficient data such as case reports, publications not in English, and studies of animal subjects. Data synthesis involved standardized extraction forms. Detailed results will be presented in the subsequent results section. A narrative synthesis of the findings will also be presented. In this systematic review, we focused on analyzing trends in the literature pertaining to the use of 7-Tesla MRI in neurosurgery. Our primary objective was to provide a comprehensive and descriptive overview of research conducted within the specified time frame.

Given the nature of our research question and the scope of our analysis, we chose not to perform a formal risk of bias assessment for individual studies using QUADAS or Rob2 [7,8]. Our decision was based on the recognition that our aim was to capture and summarize the breadth of research in this field rather than to make judgments about the quality or internal validity of the included studies nor the determination of diagnostic accuracy. Nonetheless, we have reported key characteristics and methodological details of the studies in our analysis, including any notable limitations or methodological considerations, to provide readers with a transparent understanding of the included literature.

## 3. Results

We initially identified a total of 83 records from the PubMed database between our defined period. We found no duplicate publications or any other reason to remove any manuscript before screening. All 83 articles were screened and out of them 31 were excluded as they did not meet our inclusion criteria. Finally, 52 reports were retrieved and 1 out of them was irrelevant to neurosurgery, and 3 were animal studies. This yielded a total of 48 studies that were included in our systematic review.

**Figure 1.**
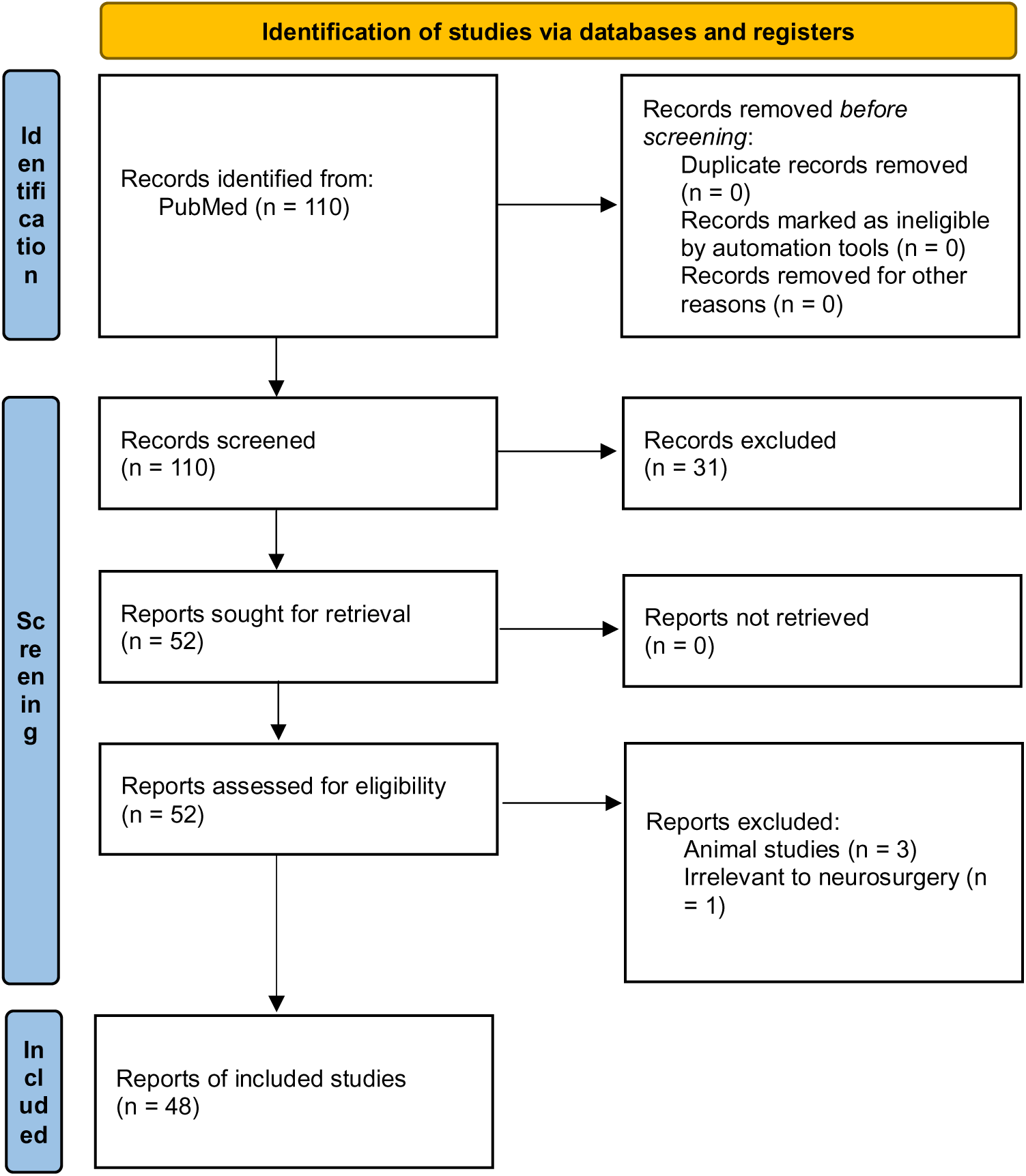
The PRISMA flowchart depicting the selection of studies for our systematic review [9].

Unfortunately, we could not conduct a meta-analysis to determine the diagnostic accuracy of the 7-T MRI for each pathology since the number of studies for each topic was deemed insufficient to derive valid conclusions.

Thus, we manually categorized the data in our database to obtain the number of publications available on PubMed for each of the neurosurgical procedures discussed in them as well as for each neurosurgically treated disease. According to our data, the most common neurosurgical procedures utilizing 7-T MRI were endoscopic neurosurgery, resective epilepsy surgery, and deep brain stimulation surgery with 5 publications (p) per each procedure, see Figure 2.

**Figure 2.**
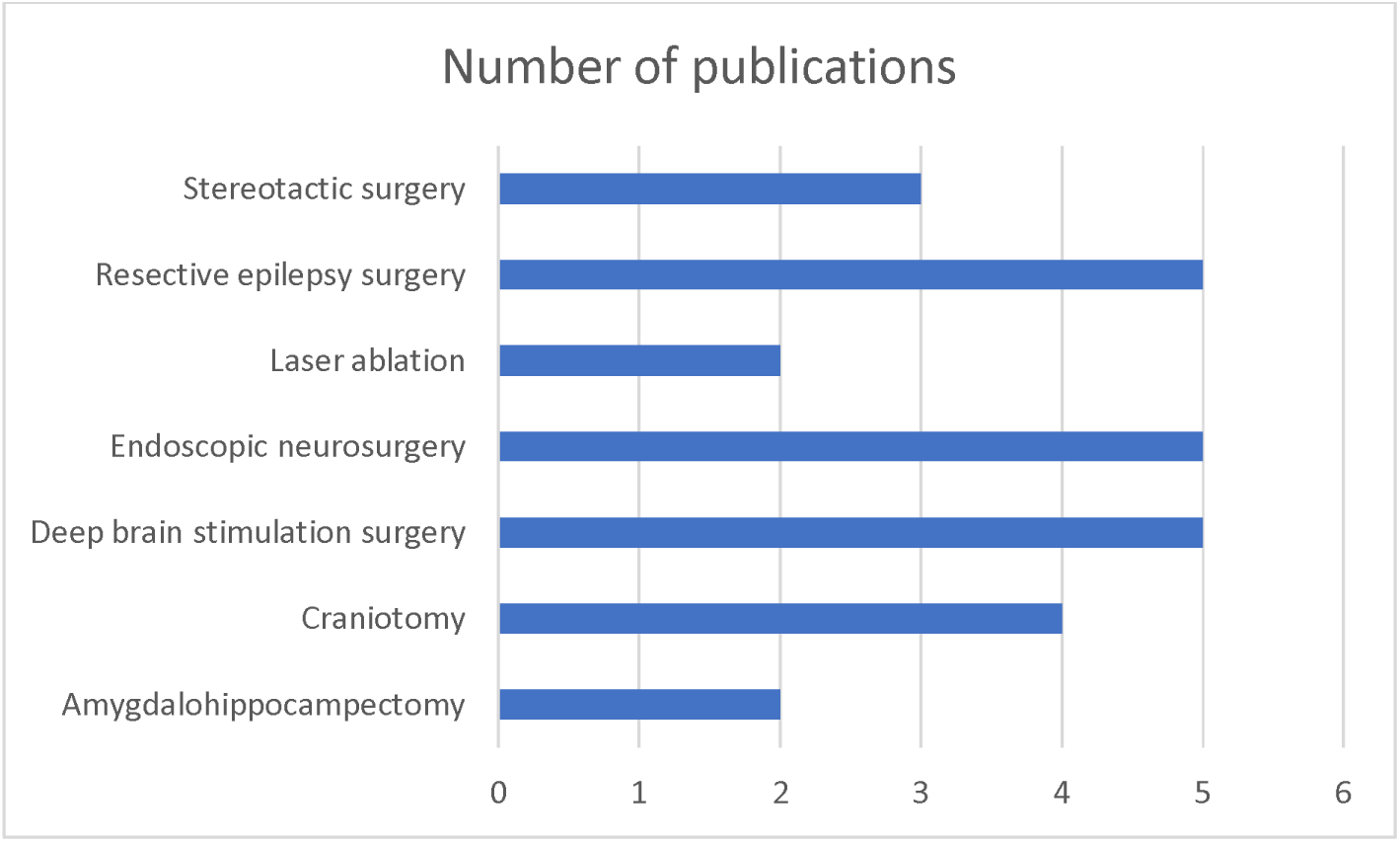
Chart showing number of 7-T publications per neurosurgical procedure.

The most commonly treated pathologies were cerebrovascular diseases (9p), followed by epilepsy (7p), pituitary adenoma (6p) and gliomas (6p). However, it should be noted that under cerebrovascular diseases, a series of diseases were considered such as amyloid angiopathy (1p), arteriovenous malformations (1p), stroke (2p), atherosclerosis (1p), intracranial aneurysms (3p), intracerebral hemorrhage (1p), see Figure 3.

**Figure 3.**
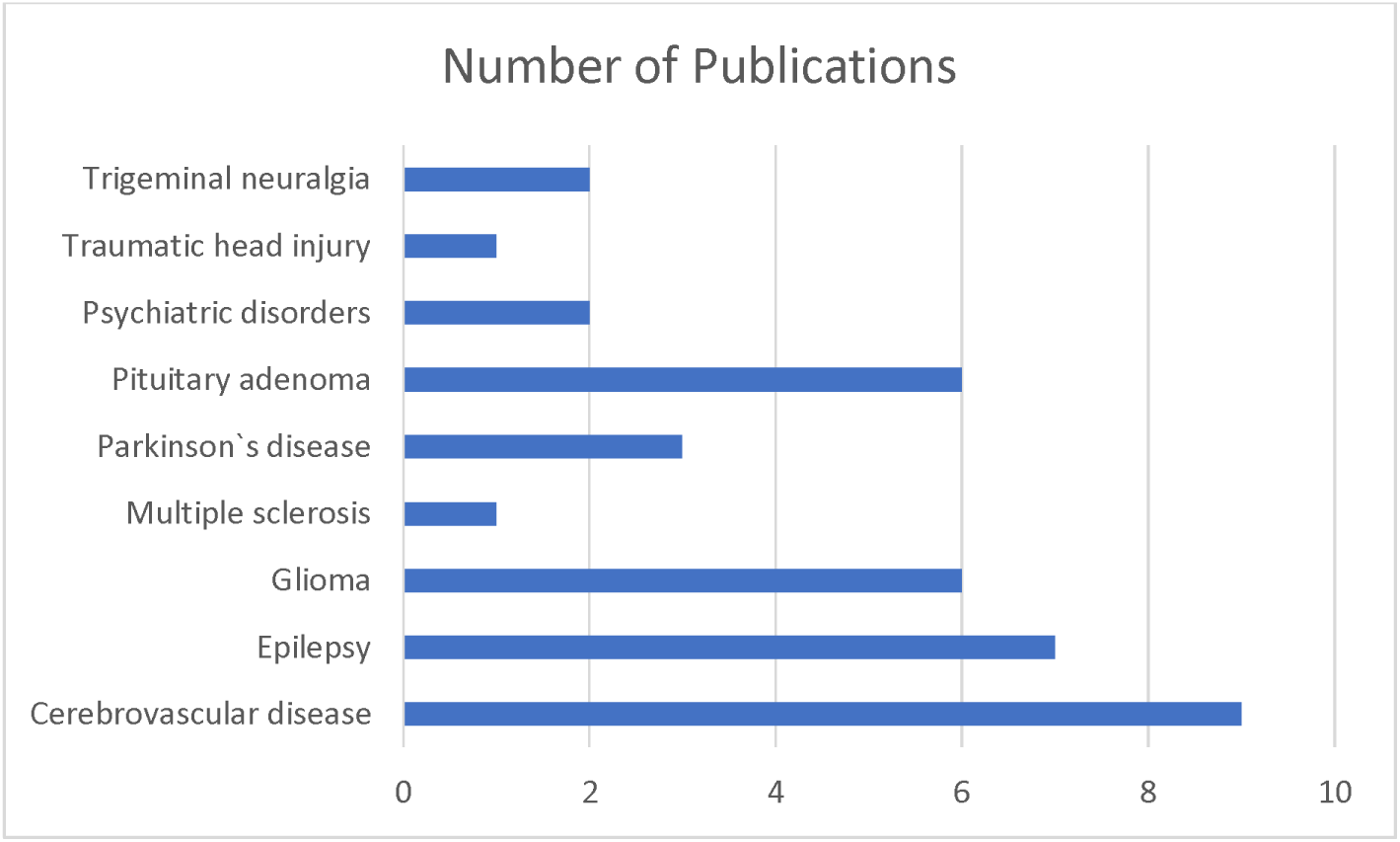
Chart showing number of 7-T publications per each neurosurgically treated pathology.

We also sorted the number of publications produced by each country. For this, only the institutional affiliation of the first author was considered. The countries with the highest absolute number of publications were the USA (15p) followed by the Netherlands (10p) and Germany (6p). However, since these values do not provide a measure of research productivity, we decided to obtain the number of publications per 7-T MRI available and concluded that the Netherlands had the highest research productivity in the present domain, see Figure 4.

**Figure 4.**
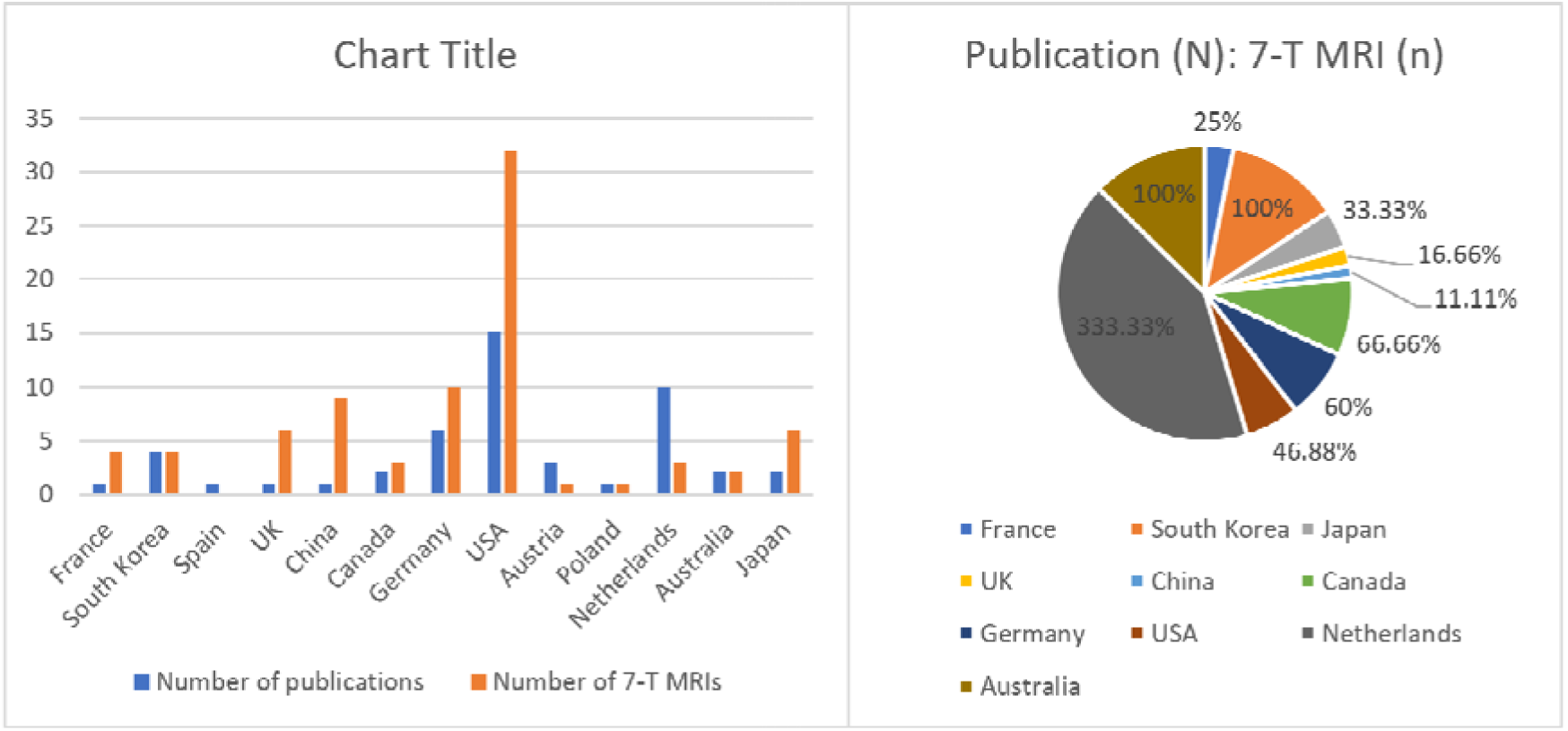
(left) Hitogram showing number of 7-T publications vs 7-T MRI machines available per country. (right) The same information translated into a pie chart to denote each country’s overall contribution.

We also noticed that there has been a decline in the research output in the year 2018 and 2021 before the subsequent increase observed later on, see Figure 5.

**Figure 5.**
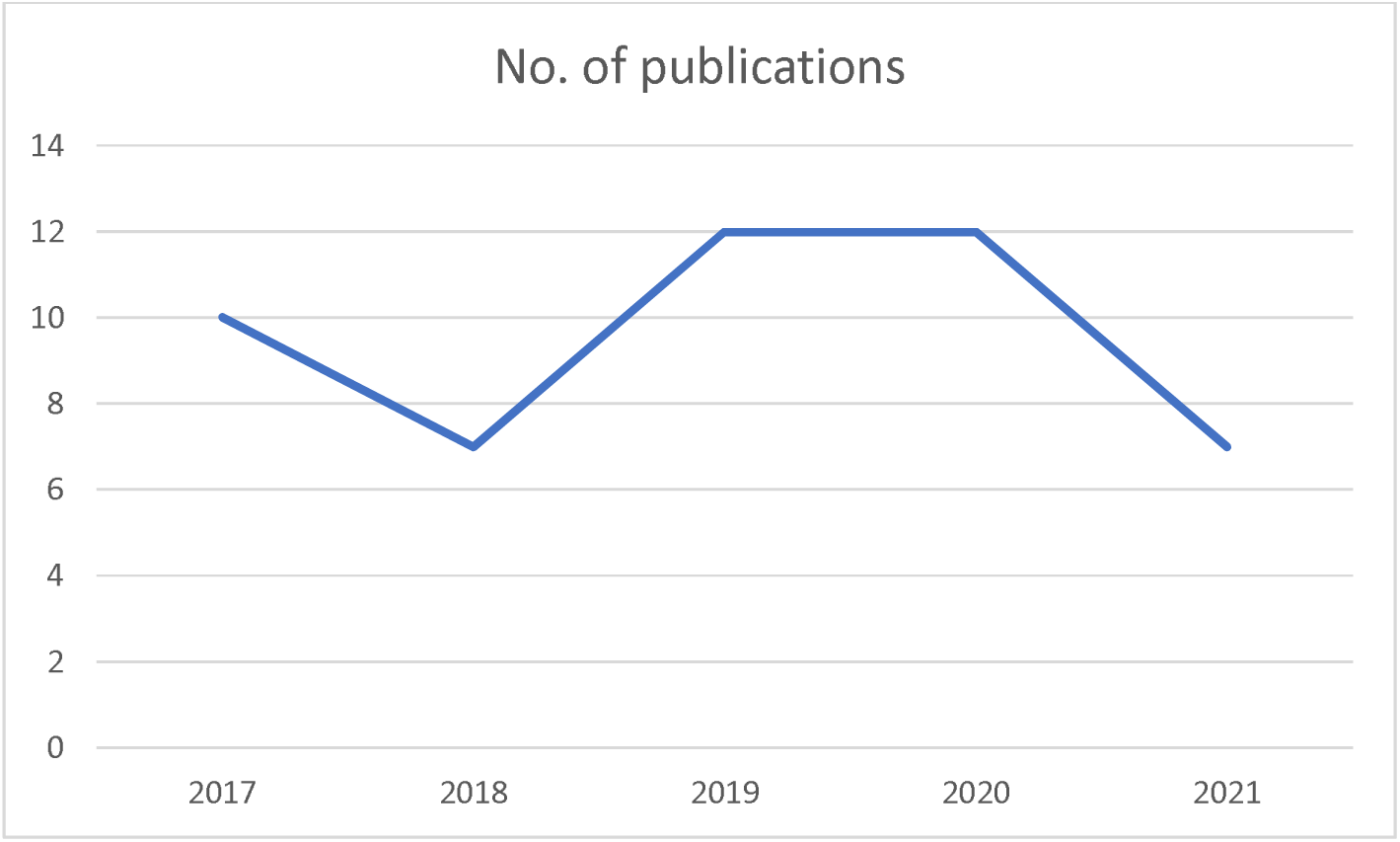
Illustration of the number of 7-T publications in each year from 2017 to 2021.

We also extracted the data from the database to create a table summarizing the studies and their primary endpoints and sequences used falling under each pathology discussed, see Table 1.

**Figure 6.**
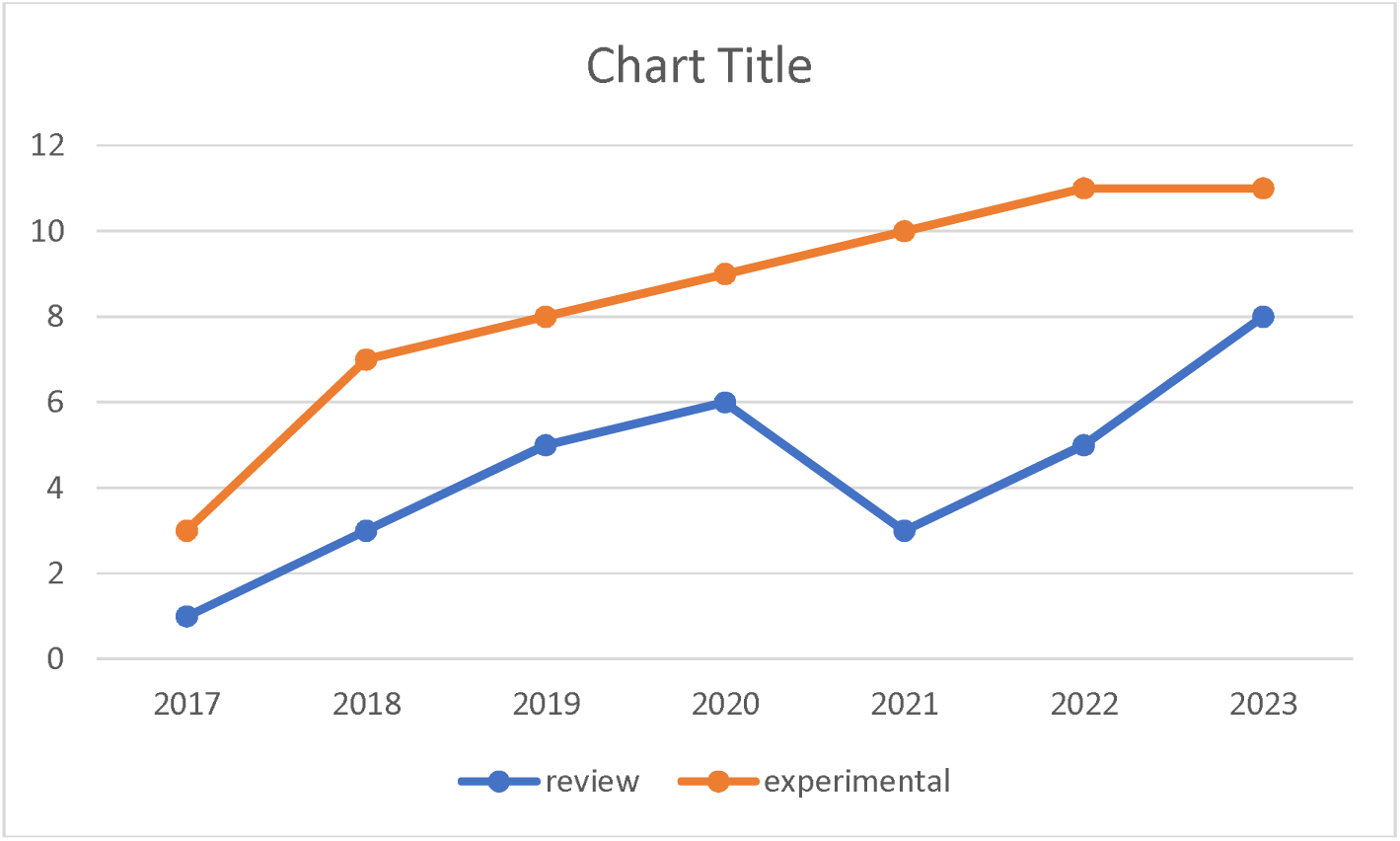
SAMPLE FIGURE !!

**Table 1:**
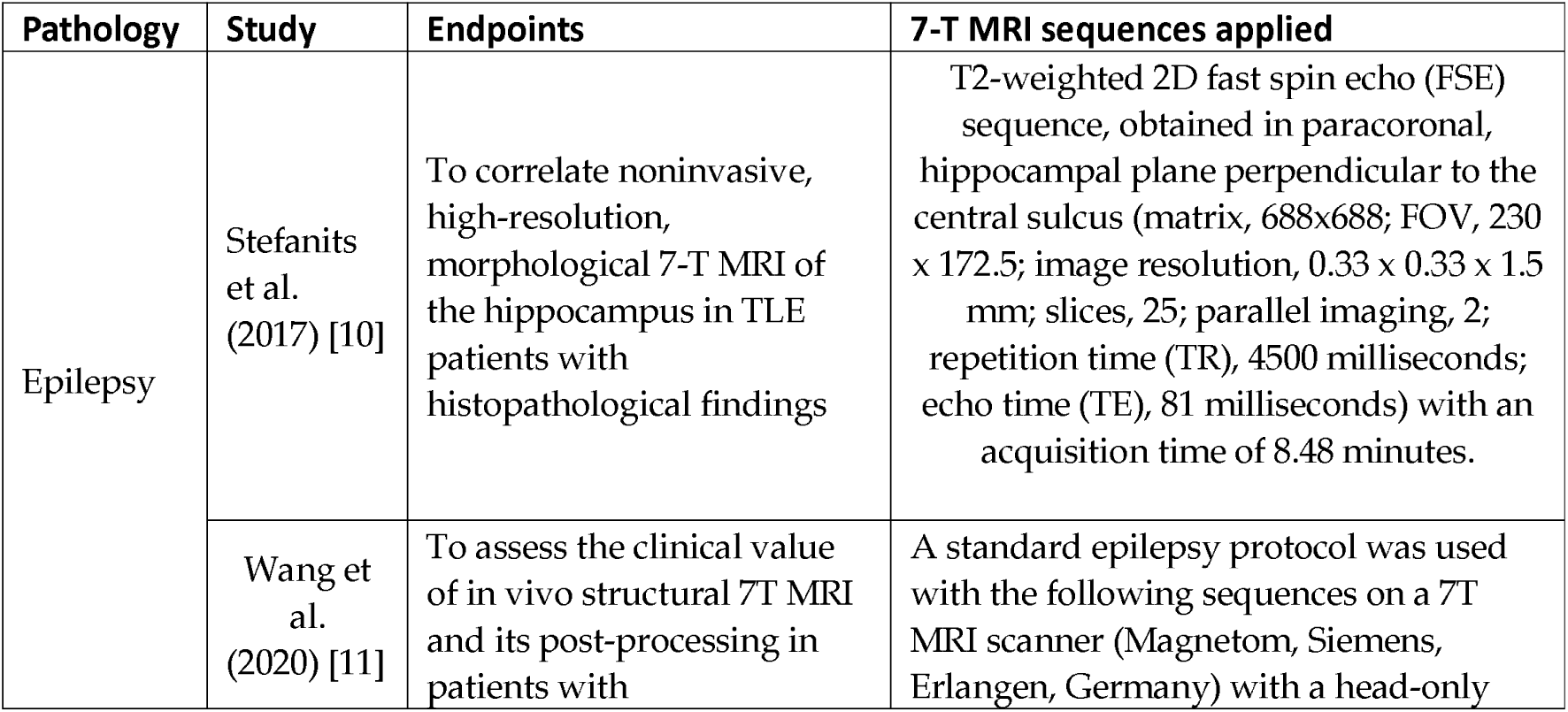
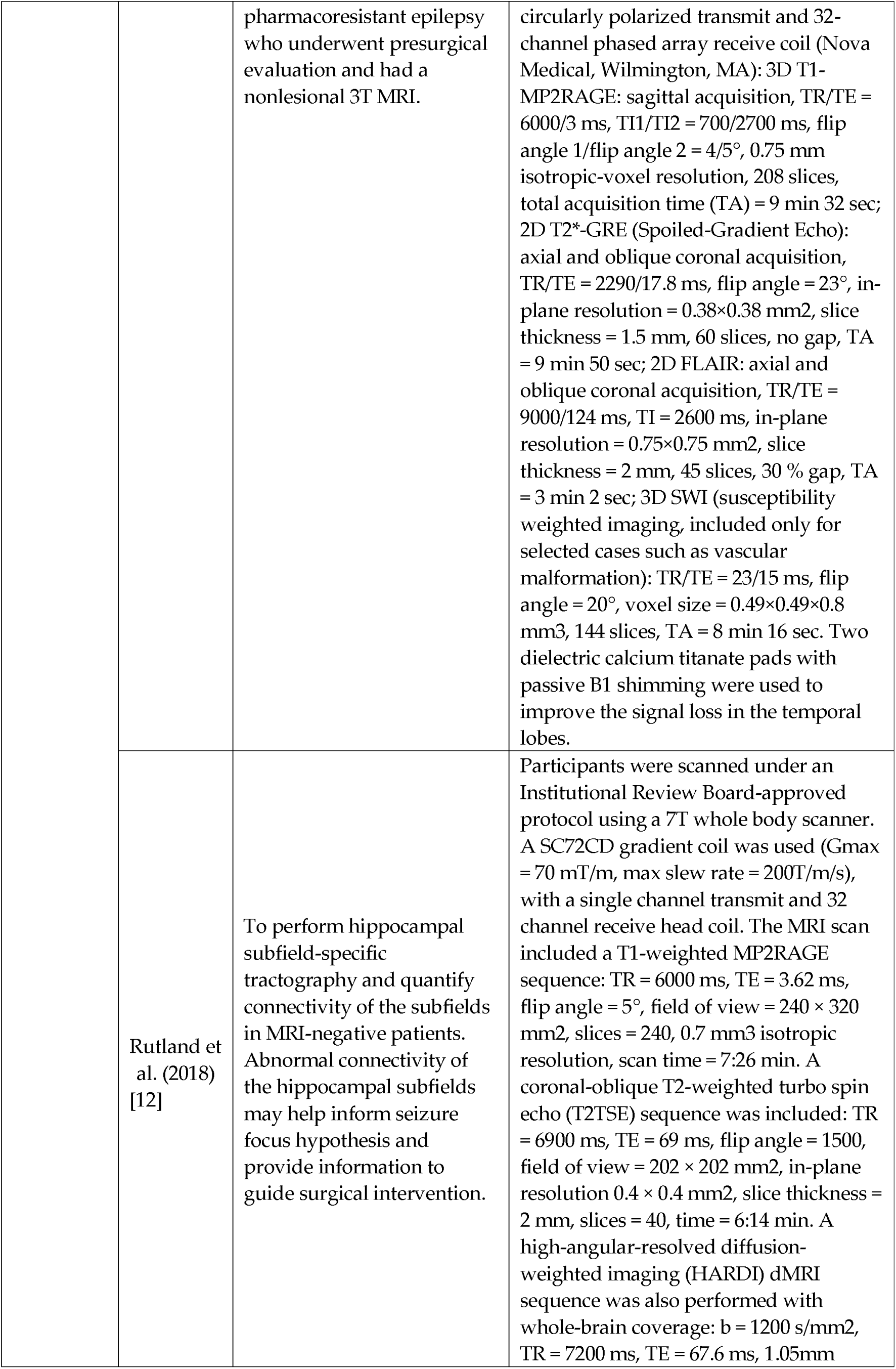
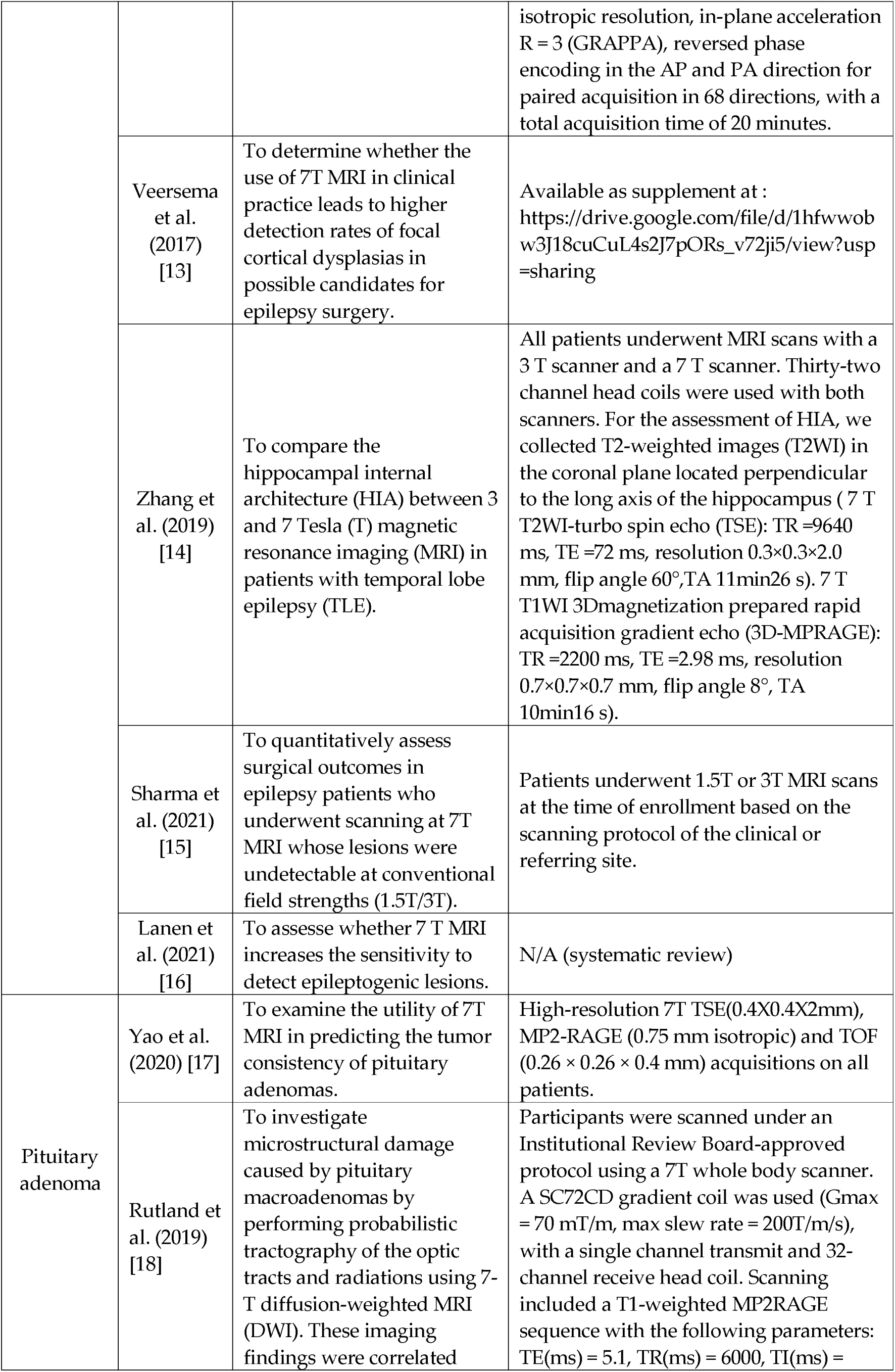
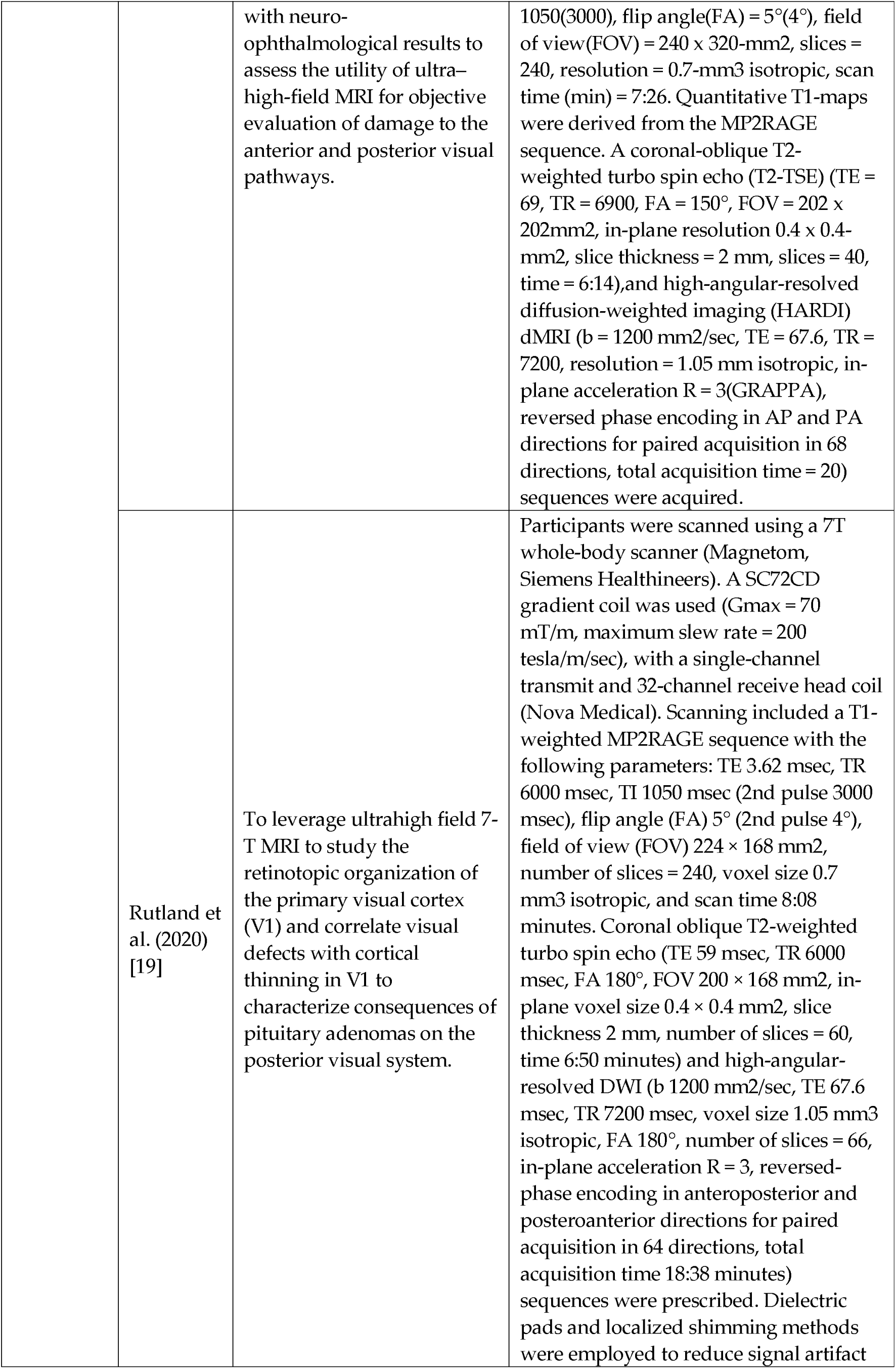
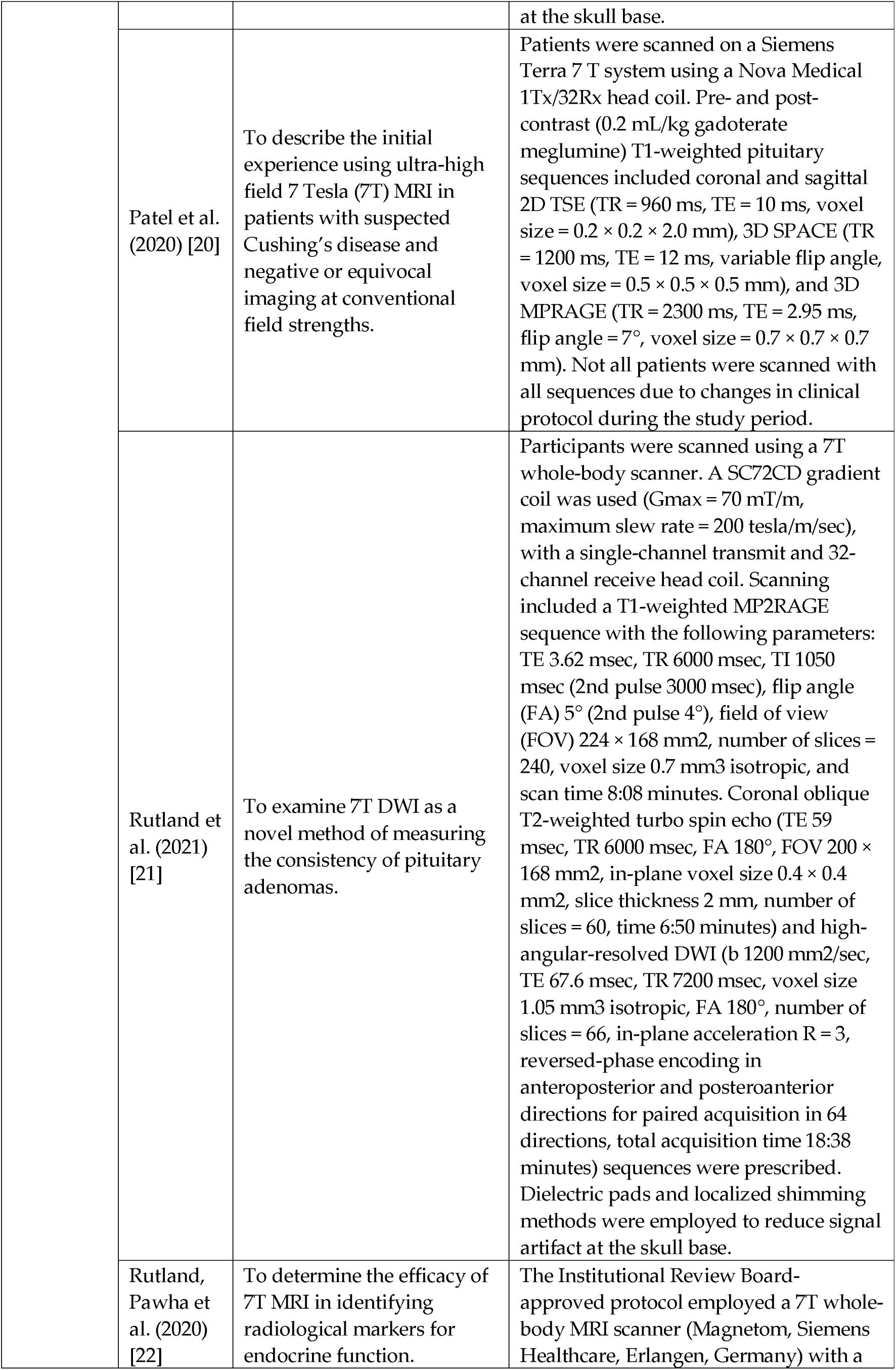
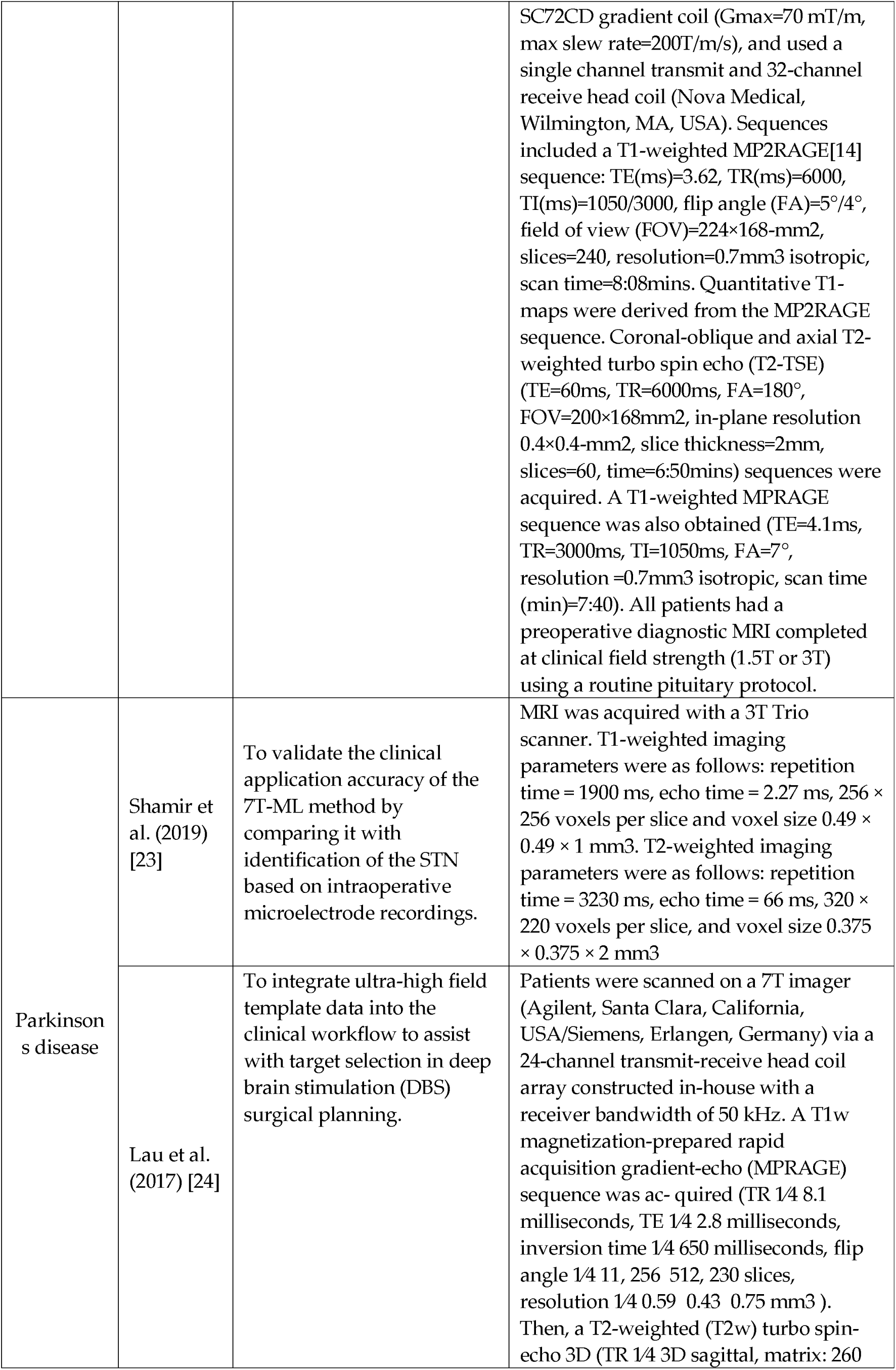
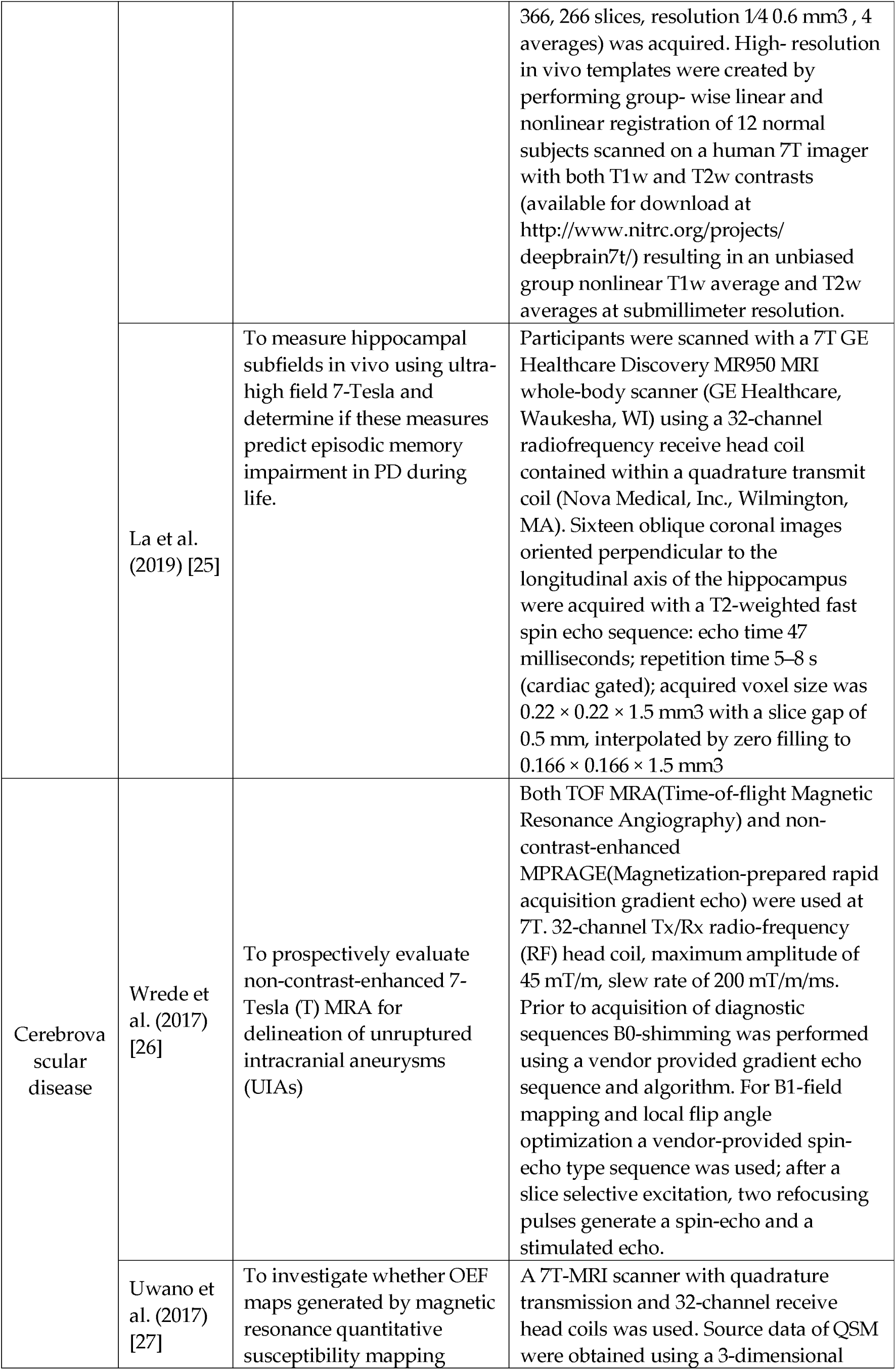
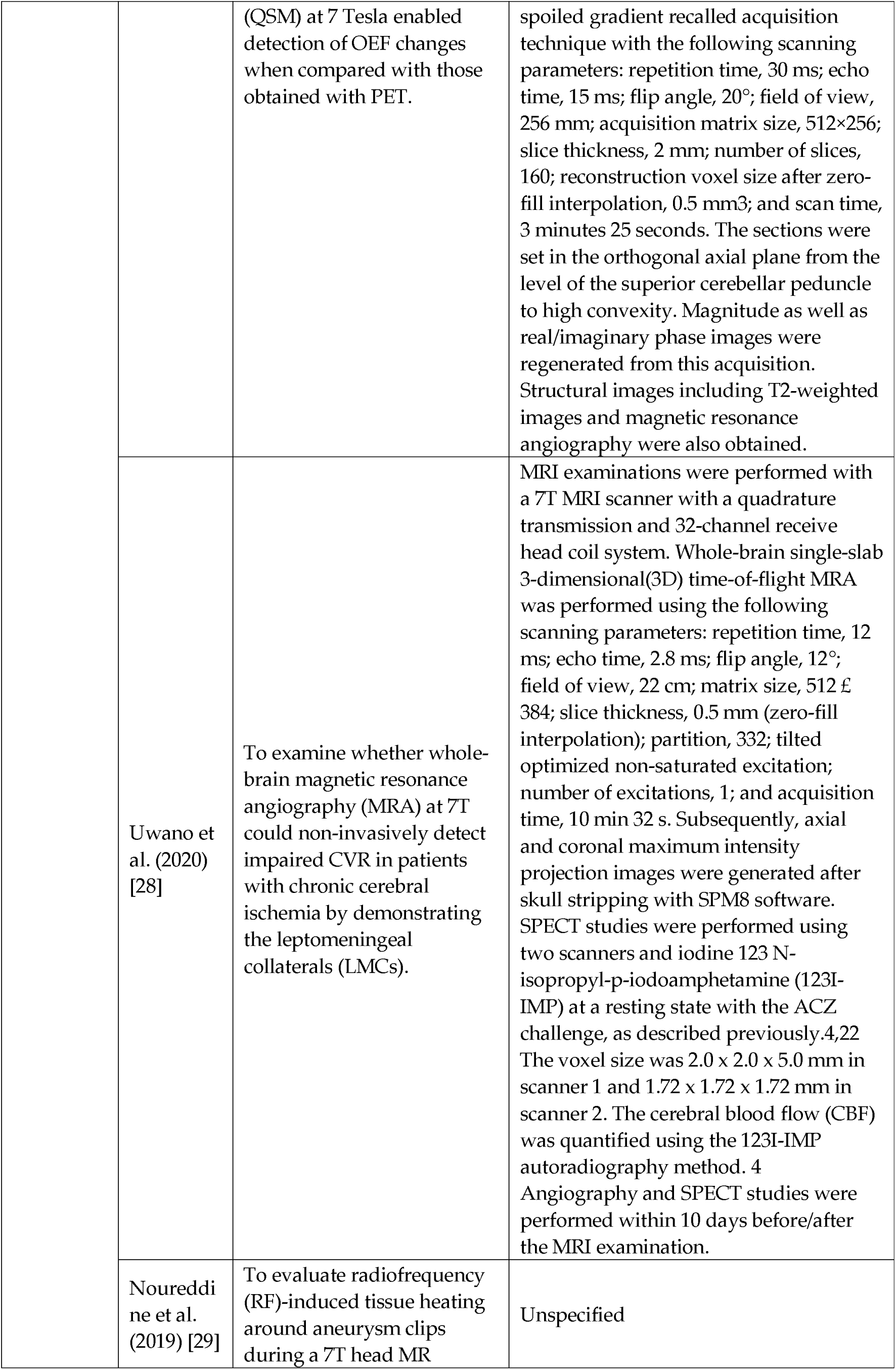
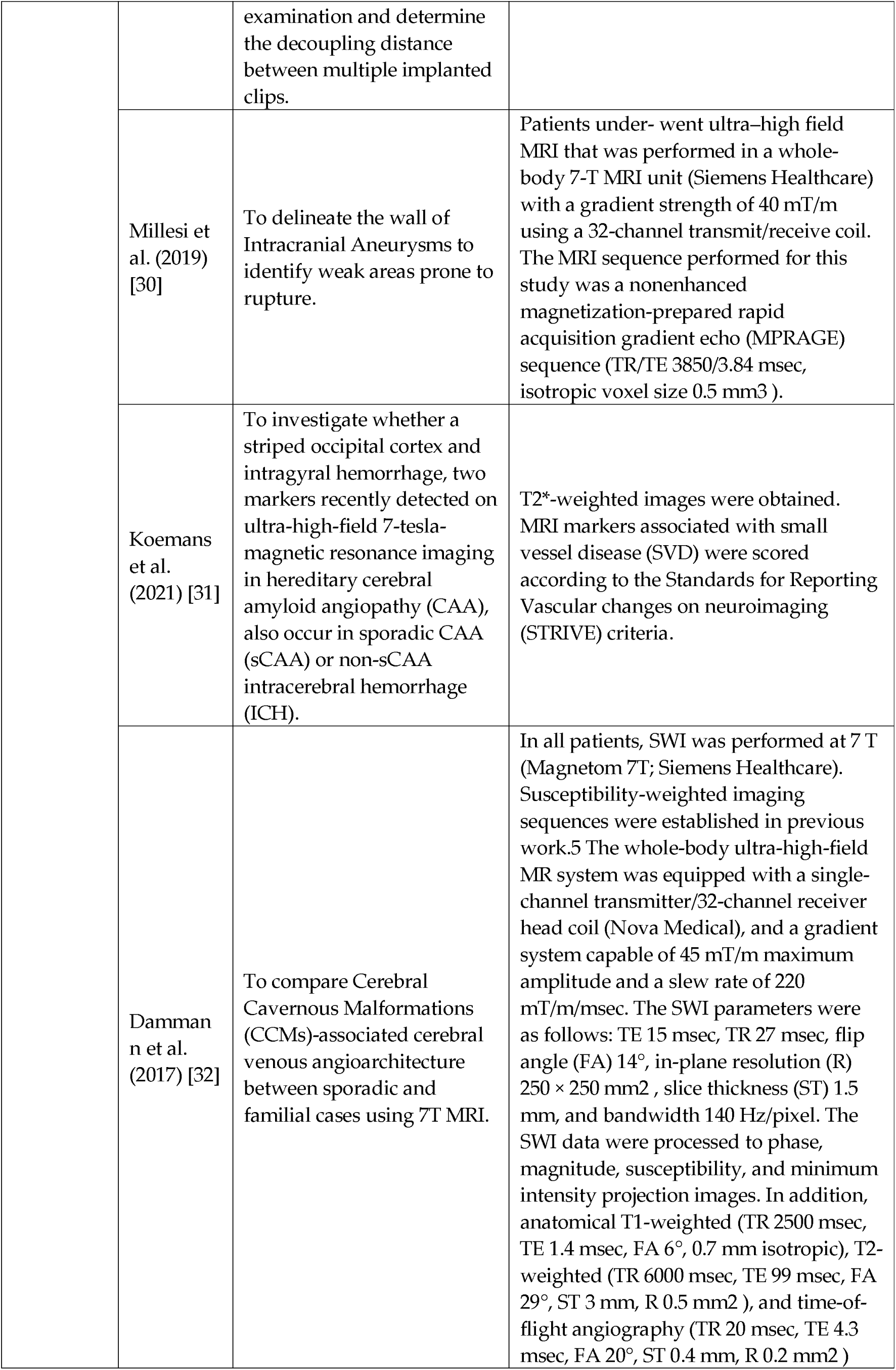
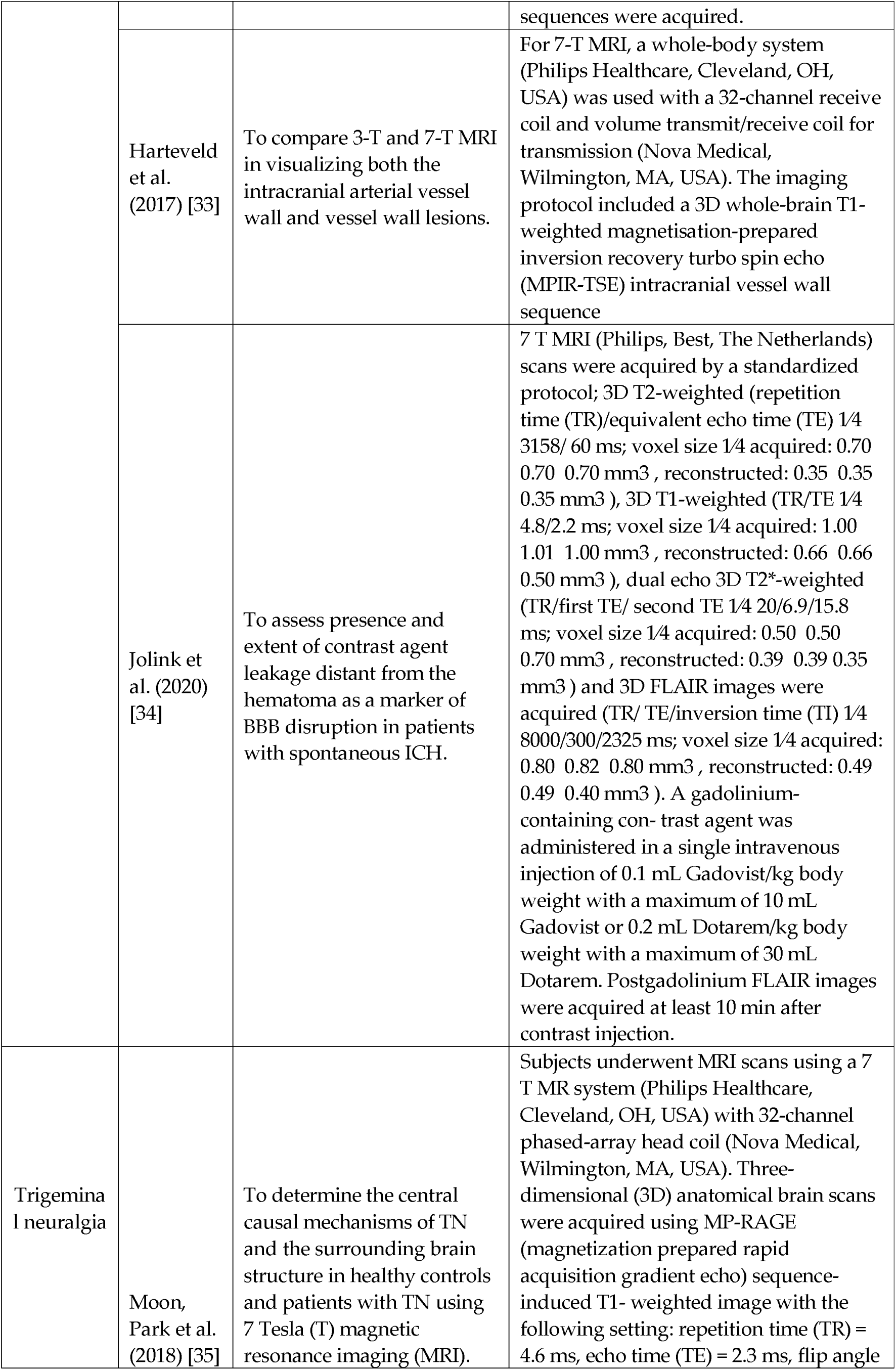
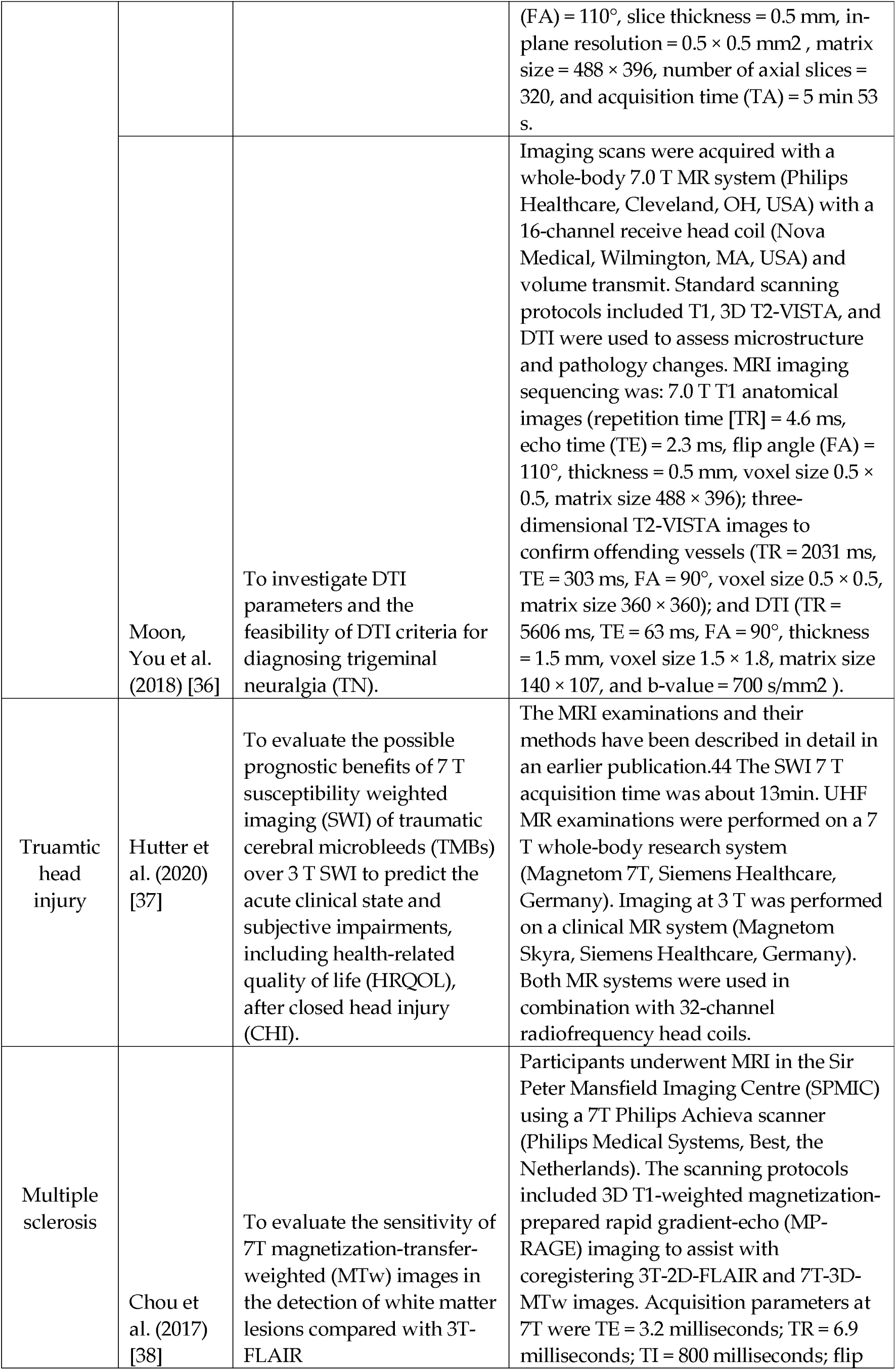
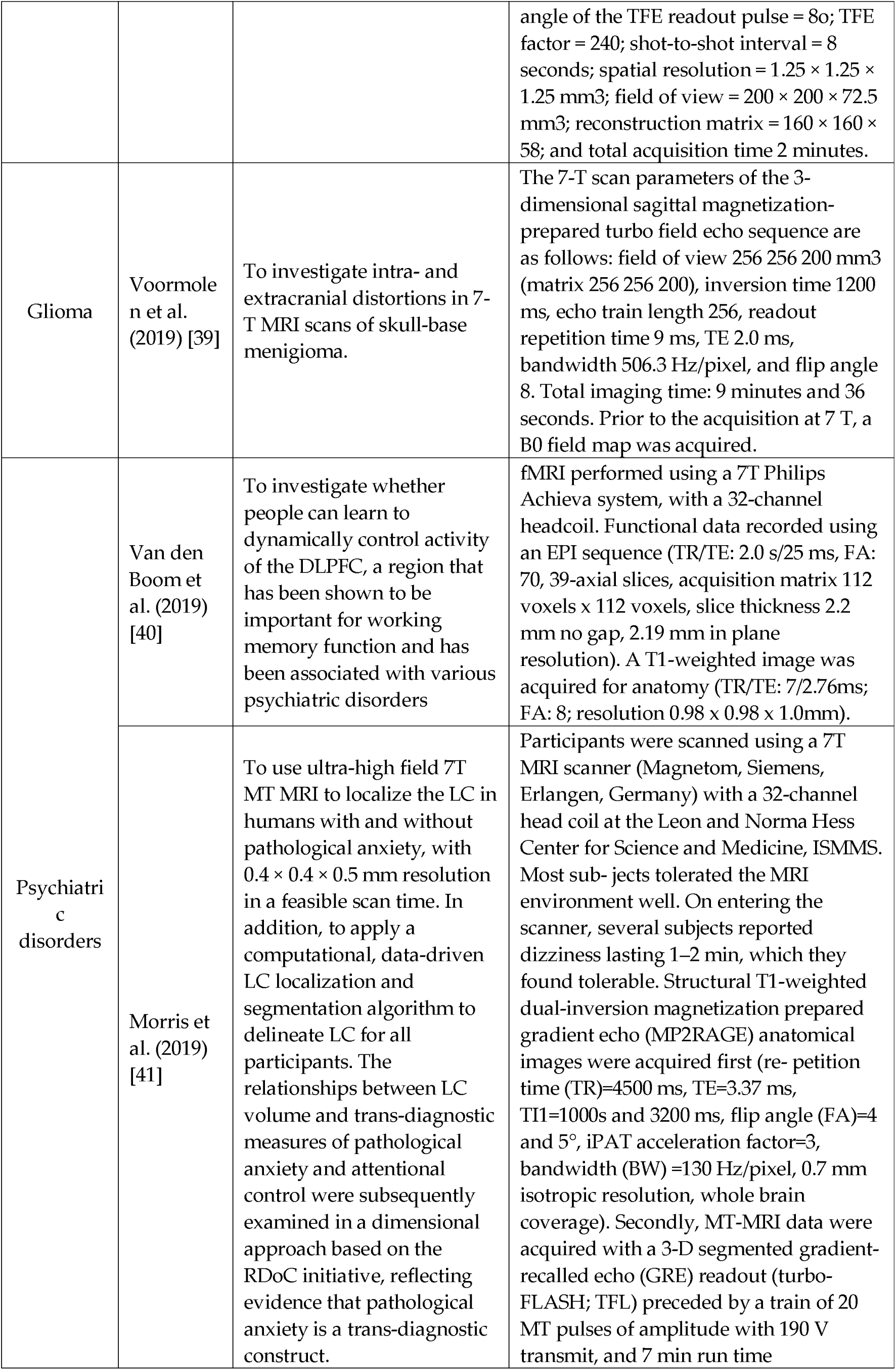
Summary of studies included in the review falling under each neurosurgically treated pathology, first author, year of publication, endpoints, and 7-T MRI protocols applied.

## 4. Discussion

The present systematic review aimed to synthesize the findings of various studies related to the use of 7T MRI in different clinical conditions. Through the analysis of the included articles, several important observations and implications have emerged. Firstly, the use of 7T MRI has demonstrated great potential in improving the characterization and understanding of various neurological and psychiatric conditions. Studies focusing on conditions such as temporal lobe epilepsy, pituitary adenoma, unruptured intracranial aneurysms, and movement disorders have reported valuable insights into disease mechanisms, anatomical abnormalities, and functional alterations. The high resolution and improved signal-to-noise ratio offered by 7T MRI have allowed for the detection of subtle structural changes and enhanced visualization of pathological features, leading to more accurate diagnosis and better treatment planning. Furthermore, the comparison studies included in this systematic review consistently demonstrated the superiority of 7T MRI over lower field strengths, such as 3T or 1.5T, in terms of image quality, lesion detection, and tissue characterization. The higher magnetic field strength of 7T MRI enables increased spatial resolution and improved contrast, which are crucial for identifying small lesions, delineating fine anatomical structures, and providing more precise localization of abnormalities.

### 4.1. Epilepsy

The utilization of high-resolution 7-T MRI in the examination of the hippocampus among Temporal Lobe Epilepsy (TLE) patients has proven to be a valuable non-invasive tool. The correlation between 7-T MRI morphological images and histopathological findings provides a crucial link, offering insights into the structural aspects of the hippocampus in TLE [10]. Overall, high-resolution, ultra-high-field MRI shows promise for detecting subtle hippocampal changes in temporal lobe epilepsy, but larger cohorts are needed to confirm its predictive value in the preoperative evaluation of TLE patients [10].

In patients with pharmacoresistant epilepsy, the inclusion of 7T MRI in the presurgical evaluation process, especially for patients with nonlesional 3T MRI scans, demonstrates its potential to uncover crucial information that may guide treatment decisions in this challenging patient population [11]. The data indicate a significant advantage of using 7T MRI with post-processing techniques for identifying subtle focal cortical dysplasia (FCD) lesions in patients with pharmacoresistant epilepsy and nonlesional 3T MRI scans. This highlights the potential of 7T imaging as a valuable tool in improving the detection of FCD lesions in challenging cases [11].

The introduction of hippocampal subfield-specific tractography has enabled the quantification of connectivity within the subfields in MRI-negative patients and identified abnormal connectivity patterns. This information holds promise in refining the understanding of seizure focus hypothesis, thereby aiding in the formulation of more informed surgical strategies [12]. The findings indicate that distinct connectivity patterns exist among hippocampal subfields in different types of epilepsy. These results hold potential significance for informing hypotheses regarding seizure focus and guiding surgical interventions, particularly in patients with MRI-negative findings. The study suggests that employing high-resolution diffusion MRI-based tractography of hippocampal subfields can reveal subtle abnormalities in patients with otherwise normal-appearing MRI scans, offering a valuable diagnostic tool in such cases [12].

7T MRI has been compared with conventional field strengths in detecting focal cortical dysplasias (FCDs). Findings suggest that the use of 7T MRI in clinical practice may significantly enhance the detection rates of FCDs, making it a valuable tool in identifying candidates for epilepsy surgery who might have otherwise been overlooked [13]. Seven-tesla MRI enhances the identification of subtle focal cortical dysplasia and mild cortical development malformations in individuals with intractable epilepsy. This improvement in detection may play a role in identifying suitable candidates for surgery and facilitating the complete resection of epileptogenic lesions, potentially leading to postoperative freedom from seizures [13].

The comparative analysis of hippocampal internal architecture between 3 and 7 Tesla MRI in TLE patients provided valuable insights into the resolution capabilities of these imaging techniques and highlighted the potential superiority of 7T MRI in capturing finer details of the hippocampal structure, contributing to a more comprehensive understanding of TLE [14]. Enhancement of hippocampal internal architecture (HIA) visualization is achievable with 7T MRI. Notably, HIA asymmetry serves as a substantial predictor for the laterality of seizure onset in temporal lobe epilepsy (TLE) patients, demonstrating comparable predictive efficacy to hippocampal volume asymmetry. Nevertheless, the utilization of 7T MRI for HIA asymmetry does not provide additional value in determining epilepsy lateralization, and it does not predict surgical outcomes [14].

The inclusion of a quantitative assessment of surgical outcomes in patients scanned at 7T MRI, whose lesions were undetectable at conventional field strengths, further emphasizes the clinical impact of 7T imaging elucidating the practical implications of employing 7T MRI in improving surgical outcomes for patients with initially elusive lesions [15]. Among the 16 patients studied, 7 exhibited clear epileptogenic potential on 7T imaging, while 9 had findings of a less definite nature. Remarkably, 15 out of 16 patients achieved Engel I, II, or III outcomes, denoting substantial improvement. Notably, those with definite lesions on 7T imaging had a higher rate of achieving Engel I surgical outcomes (57.1%) compared to those with less definite lesion status (33.3%). This suggests that patients initially diagnosed as “MRI Negative” on lower field strength scans but with clear radiological findings on 7T, corresponding to the suspected seizure onset zone (sSOZ), may benefit significantly from surgical intervention [15].

The data presented in many studies strongly suggest that the use of 7T MRI may indeed lead to increased sensitivity, thereby offering a more comprehensive diagnostic approach in the management of epilepsy [16]. The use of Ultra-High Field (UHF) MRI demonstrates increased sensitivity in detecting epileptogenic lesions, albeit with variability, indicating its potential for clinical applications. However, it remains to be determined whether this heightened sensitivity translates into improved seizure outcomes following surgical treatment. Prospective studies involving larger cohorts of epilepsy patients, consistent scan and sequence protocols, and advancements in post-processing technology are crucial for further exploration. Beyond technical enhancements, establishing a better correlation between imaging features and clinical semiology, histopathology, and overall clinical outcomes is equally important for the continued refinement of UHF MRI in epilepsy diagnosis and treatment [16].

### 4.2. Pituitary Adenoma

Our investigation into the utility of 7T MRI in predicting the tumor consistency of pituitary adenomas reveals promising insights. The high-resolution imaging capabilities of 7T MRI contribute to a more accurate characterization of the internal composition of these tumors, providing valuable information for preoperative planning and predicting surgical outcomes [17]. The enhanced signal and contrast capabilities of 7T MRI offer valuable insights into the consistency and physiology of pituitary tumors preoperatively. Employing a granular, voxel-based analysis maximizes the potential of 7T imaging resolution and represents a valuable method for predicting the consistency of pituitary adenomas [17].

7T diffusion-weighted MRI (DWI) has been successfully applied for probabilistic tractography of the optic tracts and radiations in patients with pituitary macroadenomas. The correlation between imaging findings and neuro-ophthalmological results offers a comprehensive understanding of microstructural damage. This approach contributes to the objective evaluation of damage to the anterior and posterior visual pathways, enhancing our ability to assess the impact of pituitary adenomas on visual function [18]. Quantifying secondary neuronal damage from adenomas through imaging strongly correlates with neuro-ophthalmological findings. The diffusion characteristics facilitated by ultra-high-field Diffusion-Weighted Imaging (DWI) enable preoperative characterization of visual pathway damage in patients experiencing chiasmatic compression. This approach holds the potential to inform prognosis regarding the recoverability of vision in these individuals [18].

A study also delved into the retinotopic organization of the primary visual cortex (V1) using ultra-high-field 7T MRI. By correlating visual defects with cortical thinning in V1, researchers characterized the consequences of pituitary adenomas on the posterior visual system. This exploration provides valuable insights into the anatomical basis of visual impairments associated with these tumors [19]. All 8 patients showed significant positive correlations between V1 thickness and visual defect. These findings provide retinotopic maps of localized V1 cortical neurodegeneration spatially corresponding to impairments in the visual field. These results further characterize changes in the posterior visual pathway associated with chiasmatic compression, and may prove useful in the neuroophthalmological workup for patients with pituitary macroadenoma [19].

The initial experience with ultra-high-field 7T MRI in patients with suspected Cushing’s disease and negative or equivocal imaging at conventional field strengths represents a critical aspect in detecting subtle abnormalities may offer a valuable diagnostic tool in cases where conventional imaging falls short, contributing to improved diagnostic accuracy and patient management [20]. The study revealed that 7T MRI facilitated the identification of previously unnoticed focal pituitary hypoenhancement in 90% (9/10) of patients. Remarkably, 7 out of these 9 cases corresponded histologically to corticotroph adenomas. These initial findings indicate a significant adjunctive role for ultra-high field MR imaging in the noninvasive clinical assessment of suspected Cushing’s disease [20].

7T DWI has been described as a novel method for measuring the consistency of pituitary adenomas. The high spatial resolution and sensitivity of 7T MRI enable a more detailed assessment of the internal architecture of these tumors. This novel approach holds potential for refining our understanding of tumor characteristics and guiding treatment decisions [21]. The findings from the study indicate that a high-resolution Apparent Diffusion Coefficient (ADC) of pituitary adenomas serves as a sensitive measure of tumor consistency. Utilizing 7T Diffusion-Weighted Imaging (DWI) may have clinical significance in the preoperative assessment and surgical management of patients with pituitary macroadenomas [21].

The efficacy of 7T MRI in identifying radiological markers associated with endocrine function in pituitary adenomas has been also explored. The high-field strength of 7T MRI allows for improved visualization and characterization of subtle structural changes that may be indicative of endocrine dysfunction. This contributes to a more comprehensive assessment of pituitary adenomas and their impact on hormonal regulation [22]. Radiological characterization of pituitary adenomas and the adjacent native pituitary tissue stands to benefit from the application of 7T MRI. The Corrected T2 Signal Intensity (SI) of the tumor emerges as a sensitive predictor of hormonal secretion, offering utility in the diagnostic workup for pituitary adenoma. The use of 7T MRI proves valuable in identifying markers of endocrine function in patients with pituitary adenomas. The results of the study suggest that hormone-secreting tumors exhibit higher T2-weighted SI, and tumors associated with preoperative hypopituitarism display greater stalk curvature angles [22].

### 4.3. Parkinson’s disease

A study aimed to validate the clinical application accuracy of the 7T-ML method by comparing it with the identification of the Subthalamic Nucleus (STN) based on intraoperative microelectrode recordings. The comparison highlights the reliability and precision of the 7T-ML method, suggesting its potential as a non-invasive tool for STN localization in Parkinson’s disease patients undergoing deep brain stimulation (DBS) [23]. The 7T-ML method demonstrates high consistency with microelectrode-recordings data, offering a reliable and accurate patient-specific prediction for targeting the Subthalamic Nucleus (STN) [23].

Next, the integration of ultra-high field template data into the clinical workflow represents a significant advancement in deep brain stimulation (DBS) surgical planning. By assisting with target selection, 7T MRI provides a more detailed and accurate representation of the brain anatomy, potentially improving the precision and efficacy of DBS procedures in Parkinson’s disease patients [24]. This work outlined a workflow for integrating high-resolution in vivo ultra-high field templates into the surgical navigation system to aid in Deep Brain Stimulation (DBS) planning. Importantly, this method does not impose any additional cost or time on the patient. Future efforts will focus on prospectively evaluating various templates and assessing their impact on target selection [24].

Another study delved into the in vivo measurement of hippocampal subfields using ultra-high field 7-Tesla MRI. The exploration of whether these measures predict episodic memory impairment in Parkinson’s disease during life is a novel approach. The findings provided insights into the structural changes within the hippocampus, contributing to our understanding of cognitive deficits in Parkinson’s disease and potentially offering markers for early diagnosis and intervention [25]. The study identified that the thickness of the hippocampal CA1-SP subfield, estimated from 7-Tesla MRI scans, emerged as the most robust predictor of episodic memory impairment. This predictive capability persisted even when accounting for potential confounding clinical measures. These findings suggest that ultra-high field imaging holds promise as a sensitive tool for detecting alterations in hippocampal subfields, offering insights into the neuroanatomical basis of episodic memory impairments in individuals with PD [25].

### 4.4. Cerebrovascular diseases

A prospective evaluation of the utility of non-contrast-enhanced 7-Tesla MRA for delineating unruptured intracranial aneurysms (UIAs) provided high-resolution imaging, potentially enhancing the detection and characterization of UIAs, thereby contributing to improved diagnostic capabilities in the context of cerebrovascular diseases [26]. The study showcases the outstanding delineation of Unruptured Intracranial Aneurysms (UIAs) through 7-Tesla Magnetic Resonance Angiography (MRA) within a clinical setting, presenting a comparability to the gold standard, Digital Subtraction Angiography (DSA). The combined use of 7-T non-enhanced Magnetization-Prepared Rapid Gradient Echo (MPRAGE) and Time-of-Flight (TOF) MRA for evaluating untreated UIAs emerges as a promising clinical application of ultra-high-field MRA [26].

The potential of OEF maps generated by magnetic resonance quantitative susceptibility mapping (QSM) at 7 Tesla in detecting changes in oxygen extraction fraction (OEF) has also been investigated. The comparison with PET results establishes the capability of 7T MRI to non-invasively monitor OEF alterations, offering a valuable alternative to traditional imaging modalities [27]. In patients with unilateral steno-occlusive internal carotid artery/middle cerebral artery lesions, OEF ratios on 7-Tesla

Quantitative Susceptibility Mapping (QSM) images demonstrated a strong correlation with those on Positron Emission Tomography (PET) images. This suggests that noninvasive OEF measurement by MRI has the potential to serve as a substitute for PET in assessing oxygen extraction fraction [27].

A study also explored the use of whole-brain magnetic resonance angiography (MRA) at 7T for the non-invasive detection of impaired cerebrovascular reactivity (CVR) in patients with chronic cerebral ischemia. By highlighting leptomeningeal collaterals (LMCs), 7T MRI emerges as a promising tool for assessing cerebrovascular function in a comprehensive manner [28]. The development of LMCs on whole-brain MRA at 7T can non-invasively detect reduced CVR with a high sensitivity/specificity in patients with unilateral cervical stenosis [28].

The radiofrequency (RF)-induced tissue heating around aneurysm clips during a 7T head MR examination was also evaluated and aimed to determine the decoupling distance between multiple implanted clips, ensuring the safety and feasibility of 7T MRI in patients with aneurysm clips [29]. In a 7T ultra-high field MRI setting, safe scanning conditions regarding Radiofrequency (RF)-induced heating can be implemented for single or decoupled aneurysm clips. However, more research is required for cases involving multiple aneurysm clips separated by less than 35 mm to ensure safety in this specific configuration [29].

Delineation of wall weak areas in Intracranial Aneurysms was performed using 7T MRI to identify weak areas prone to rupture. This approach provided valuable insights into the structural characteristics of aneurysm walls, potentially contributing to risk stratification and personalized treatment strategies [30]. In most cases, there was an observed hyperintense rim effect along the vessel wall. Notably, focal irregularities within this rim demonstrated elevated values of mean wall shear stress and vorticity, as analyzed by Computational Fluid Dynamics (CFD). These findings suggest that there are alterations in blood flow within these specific areas in Intracranial Aneurysms (IAs) [30].

The occurrence of a striped occipital cortex and intragyral hemorrhage, previously detected on ultra-high-field 7-tesla magnetic resonance imaging in hereditary cerebral amyloid angiopathy (CAA) was also investigated. The study aimed to determine whether these markers are also present in sporadic CAA (sCAA) or non-sCAA intracerebral hemorrhage (ICH), contributing to the understanding of imaging markers in different forms of CAA [31]. While a striped occipital cortex is uncommon in superficial cortical siderosis (sCAA), approximately 12% of patients with sCAA exhibit intragyral hemorrhages. These intragyral hemorrhages appear to be associated with advanced disease, and their absence in patients with non-superficial cortical siderosis intracerebral hemorrhages (non-sCAA-ICH) may suggest a level of specificity for cerebral amyloid angiopathy (CAA) [31].

Next, a study compared cerebral cavernous malformations (CCMs)-associated cerebral venous angioarchitecture between sporadic and familial cases using 7T MRI. This investigation provided insights into the vascular alterations associated with CCMs, potentially aiding in the differentiation and characterization of familial and sporadic cases [32]. The SWI results of the venous angioarchitecture of multiple CCMs correlate with sporadic or familial disease. These results are consistent with the theory that venous anomalies are causative for the sporadic form of multiple CCMs [32].

A comparison of 3T and 7T MRI in visualizing intracranial arterial vessel wall and vessel wall lesions found potential benefits of 7T MRI in providing enhanced resolution and detailed characterization of vessel wall abnormalities in cerebrovascular diseases [33]. Even with considerable variability in detected lesions at both field strengths, 7-Tesla MRI has the highest potential for identifying the overall burden of intracranial vessel wall lesions [33].

The presence and extent of contrast agent leakage distant from the hematoma was found as a marker of blood-brain barrier (BBB) disruption in patients with spontaneous intracerebral hemorrhage (ICH). The use of 7T MRI allows for a detailed examination of BBB integrity, providing valuable information on the extent of vascular damage in the surrounding brain tissue [34]. This study shows that contrast leakage distant from the hematoma is common in days to weeks after spontaneous ICH. It is located predominantly cortical and related to lobar CMBs and therefore possibly to cerebral amyloid angiopathy [34].

### 4.5. Trigeminal neuralgia

7 Tesla MRI was employed to investigate the central causal mechanisms of trigeminal neuralgia (TN) and the surrounding brain structure. By comparing healthy controls with patients suffering from TN, researchers aimed to identify structural and functional alterations in the trigeminal nerve and associated brain regions. The high resolution and sensitivity of 7T MRI enable a detailed examination, shedding light on potential biomarkers and contributing to our understanding of the underlying mechanisms of TN [35]. Results suggest that the cACC, PCC but not the rACC are associated with central pain mechanisms in TN [35].

Another study explored the use of 7T MRI for investigating diffusion tensor imaging (DTI) parameters and assessing the feasibility of DTI criteria for diagnosing trigeminal neuralgia. By examining the microstructural integrity of the trigeminal nerve, the research aimed to identify specific DTI parameters that may serve as diagnostic indicators for TN. The high-field strength of 7T MRI enhanced the accuracy and sensitivity of DTI measurements, providing valuable information for the development of reliable diagnostic criteria for trigeminal neuralgia [36]. The heightened signal-to-noise ratio offered by 7 Tesla MRI is expected to be advantageous in enhancing spatial resolution for the detection of microstructure changes in trigeminal nerves among patients with Trigeminal Neuralgia (TN) [36].

### 4.6. Traumatic head injury

A study examined the potential prognostic advantages of utilizing 7 Tesla susceptibility-weighted imaging (SWI) for traumatic cerebral microbleeds (TMBs) in comparison to 3 Tesla SWI. This evaluation aimed to forecast the immediate clinical condition and subjective impairments, encompassing health-related quality of life (HRQOL), following a closed head injury (CHI). The number of traumatic cerebral microbleeds showed a substantial association with indicators of the acute clinical state and chronic neurobehavioral parameters after closed head injury, but there was no additional advantage of 7 T MRI. These preliminary findings warrant a larger prospective study for the future [37]. The number of TMBs showed a substantial association with indicators of the acute clinical state and chronic neurobehavioral parameters after CHI, but there was no additional advantage of 7 T MRI. These preliminary findings warrant a larger prospective study for the future [37].

### 4.7. Multiple sclerosis

Fluid-attenuated inversion recovery (FLAIR) imaging at a 3 Tesla (T) field strength is recognized as the most sensitive method for identifying white matter lesions in multiple sclerosis. Although 7T FLAIR effectively detects cortical lesions, it has not been fully optimized for visualizing white matter lesions, thus limiting its use in delineating lesions in quantitative magnetic resonance imaging (MRI) studies of normal appearing white matter in multiple sclerosis. As a result, a team assessed the sensitivity of 7T magnetization-transfer-weighted (MTw) images in detecting white matter lesions in comparison to 3T FLAIR. The findings indicate that 7T MTw sequences successfully identified a majority of white matter lesions detected by FLAIR at 3T. This implies that 7T-MTw imaging serves as a robust alternative for detecting demyelinating lesions alongside 3T FLAIR. Subsequent studies should explore and compare the roles of optimized 7T-FLAIR and 7T-MTw imaging [38]. Seven-Tesla Magnetization Transfer-weighted (MTw) sequences successfully detected the majority of white matter lesions that were identified by Fluid-Attenuated Inversion Recovery (FLAIR) at 3 Tesla. This indicates that 7T-MTw imaging can serve as a robust alternative for detecting demyelinating lesions in addition to 3T-FLAIR. Future studies should focus on comparing the roles of optimized 7T-FLAIR and 7T-MTw imaging for a more comprehensive understanding of their respective contributions [38].

### 4.8. Glioma

Ultra-high-field magnetic resonance imaging (MRI) of the brain is an appealing option for image guidance during neurosurgery due to its superior tissue contrast and detailed vessel visualization. However, the susceptibility of high-field MRI to distortion artifacts poses a potential challenge to image guidance accuracy. In this study, we specifically examine intra-and extracranial distortions in 7-T MRI scans. Upon inspection of magnetization-prepared T1-weighted 7-T MRI cranial images, no discernible intracranial distortions were observed. However, noteworthy extracranial shifts were identified. These shifts introduce a level of unreliability in 7-T images when used for patient-to-image registration. To address this issue, researchers recommend conducting patient-to-image registration on a standard imaging modality, such as a routine computed tomography scan or a 3-T magnetic resonance image. Subsequently, the 7-T MRI can be fused with the routine image on the image guidance machine. This proposed approach is advised until a resolution is achieved for the observed extracranial shifts in 7-T MRI scans [39]. Magnetization-prepared T1-weighted 7-Tesla MRI cranial images exhibit no visible intracranial distortions but significant extracranial shifts. These shifts make patient-to-image registration on 7-T images unreliable. As a solution, we recommend conducting patient-to-image registration on a routine image (such as computed tomography or 3-Tesla magnetic resonance) and then fusing the 7-T magnetic resonance image with the routine image on the image guidance machine until this issue is addressed [39].

### 4.9. Psychiatric disorders

A team explored the potential for individuals to acquire the skill of dynamically controlling the activity of the dorsolateral prefrontal cortex (DLPFC). The DLPFC is recognized for its significance in working memory function and its association with various psychiatric disorders. This study also seeked to delve into the learnability of such dynamic control, which may have implications for our understanding of cognitive processes and mental health conditions [40]. These findings offer an initial indication that individuals may have the capacity to learn to dynamically down-regulate physiological activity in the Dorsolateral Prefrontal Cortex (DLPFC). This has potential implications for psychiatric disorders in which the DLPFC plays a significant role [40].

Another study utilized a combination of high-resolution and quantitative magnetic resonance (MR) imaging, employing both supervised and unsupervised computational techniques. This approach enabled the acquisition of robust sub-millimeter measurements of the locus coeruleus (LC) in vivo.

Additionally, the study investigated the correlation of these measurements with prevalent psychopathological conditions. The implications of this work extend broadly, as it holds the potential to impact various neurological and psychiatric disorders known for their association with anticipated LC dysfunction [41]. This study combined high-resolution and quantitative MR with a mixture of supervised and unsupervised computational techniques to provide robust, sub-millimeter measurements of the LC in vivo, which were additionally related to common psychopathology. This work has wide-reaching applications for a range of neurological and psychiatric disorders characterized by expected LC dysfunction [41].

## 5. Limitations of our analysis

It is essential to highlight the limitations of our study. The first limitation is that in order to make the analysis feasible and to ensure the credibility of the studies we have only used the PubMed database since we observed that manually searching Embase there was no additional significant gain in terms of the number of studies.

Furthermore, while we have evaluated the research productivity using the ratio of publications per resource, it’s important to keep in mind that research productivity should not be evaluated solely based on a single metric like this ratio. Productivity in research is multifaceted and can be influenced by various factors, including the quality and impact of publications, the significance of the research findings, collaborations, funding, and more. So, we acknowledge that this ratio should have been used in conjunction with other relevant metrics and qualitative assessments to provide a more comprehensive evaluation of research productivity in your specific context.

Next, with regard to Spain that showed 1 publication and no 7T MRI facility, we hypothesize that this was probably because they might have conducted the study in a facility outside Spain. So, due to this issue (a value of 0 in the denominator giving an infinitely high result) Austria, Poland, and Spain were removed for the sake of obtaining a valid dataset.

Another limitation of our study was that if an author affiliated to the Neurosurgery department was involved in the manuscript, the study was included even though a procedure was not specified in the article.

In addition, the reduction of number of publications in 2021 could have been due to the fact that some records might have been added to the database after our search in January 2022.

Lastly, though we retrieved a total of 48 studies into our database, only 31 out of those have been referenced in this review in Table 1 since they were relevant to a specific pathology while the remaining have only been denoted in the figures (were used only for analysis and generation of the data) [41–58].

## 6. Conclusion

The findings open avenues for further exploration and integration of 7T imaging into routine clinical practice, promising improved patient outcomes and refined surgical interventions. However, this will be possible only if the distribution of 7-T MR systems is accelerated worldwide and given that radiologists receive enough training regarding safety measures, feasibility, and other challenges.

## Data Availability

All data produced in the present study are available upon reasonable request to the authors

## Acknowledgement

The LLM ChatGPT has been used for paraphrasing [59–67].

(1) A continuously updated list of 7-T MRI facilities and their relevant locations globally can be accessed at https://www.google.com/maps/d/u/0/viewer?ll=1.941826124046989%2C0&z=2&mid=1dXG84OZIAOxjsqh3x2tGzWL1bNU

(2) The second reviewer (or author) decided to not include his name in the list of authors.

